# A Randomized Controlled Trial of Cognitive Behavioral Therapy for Insomnia During Early Recovery from Alcohol Use Disorder Among Veterans

**DOI:** 10.1101/2025.01.03.25319973

**Authors:** Subhajit Chakravorty, Knashawn H. Morales, Michael L. Perlis, Samuel T. Kuna, Sean He, David W. Oslin, J. Todd Arnedt, Henry R. Kranzler

**Author notes:** Corresponding Author: Subhajit Chakravorty, MD MIRECC, 2^nd^ Floor, Mail stop 116 Cpl. Michael J. Crescenz VA Medical Center, 3900 Woodland Avenue, Philadelphia, PA 19104.

## Abstract

**Study Objectives:** 1) To determine the efficacy of Cognitive Behavioral Therapy for Insomnia (CBT-I) for improving insomnia, alcohol-related outcomes, and daytime functioning at post-treatment and at 3- and 6- month follow-up, in a largely African American Veteran sample; 2) Evaluate whether improvement in insomnia is associated with a reduction in alcohol-related outcomes post-treatment.

**Methods:** An RCT of CBT-I (n = 31) compared to Quasi-Desensitization therapy (QDT, n = 32), eight weekly in-person sessions, with assessments at baseline, end of treatment (8 weeks), and 3- and 6-months post-treatment. Primary outcomes were the Insomnia Severity Index (ISI) total score, and Percent Days Abstinent (PDA). Secondary outcomes were sleep diary variables, drinks per day, percentage non-heavy drinking days, Penn Alcohol Craving Scale, PCS and MCS scale (from the SF-12), BDI and STAI-Trait subscale total scores.

**Results:** Post-treatment data were obtained from 88.9% of participants. Although CBT-I improved insomnia with effect sizes (E.S.) larger than the meta-analytic estimates, QDT was equally efficacious in improving insomnia (E.S. = -1.63 vs. -1.50), improving abstinence (E.S. = 1.54 vs. 1.91) and next-day functioning (E.S. = - 0.26 vs. -0.17). Across treatment groups, remission from insomnia was associated with a lower post-treatment alcohol craving score (2.79, 95% CI 1.14, 4.44 vs. 9.51, 95% CI 6.06, 12.95 in non-responders), an effect that persisted for 6 months after treatment.

**Conclusions:** CBT-I and QDT are equally effective for treating insomnia during early recovery from AUD. Reduced alcohol craving may be a mechanism by which a remission from insomnia improves drinking outcomes.

The study was registered on ClinicalTrials.gov: NCT01987089.

**Statement of Significance:** Insomnia is prevalent in alcohol use disorder (AUD) and predicts relapse during early recovery from pathological drinking. Cognitive Behavioral Therapy for Insomnia (CBT-I) is a promising treatment for this co-occurring condition. This randomized-controlled study, with a predominantly African-American sample, compared CBT-I to Quasi-Desensitization Therapy (QDT), an active placebo. Both treatments effectively improved insomnia, promoted abstinence, and enhanced next-day functioning. Importantly, across the entire sample, remission from insomnia was associated with reduced alcohol craving for up to six months after treatment. However, QDT proved equally efficacious as CBT-I in treating insomnia during early recovery from AUD. These findings suggest that while remission from insomnia may be associated with reduced alcohol craving, QDT was as efficacious as CBT-I in promoting recovery.

## Introduction

Insomnia is prevalent in 36-91% of people with Alcohol Use Disorder (AUD)^1^. Compared to individuals without insomnia, those with co-occurring insomnia and AUD have a higher severity of AUD, a poorer quality of life, greater alcohol craving, are more likely to relapse to drinking during early recovery, and are at increased risk of suicidal behavior^1^. Prior studies of medication treatments, such as gabapentin, trazodone, and quetiapine, have shown inconsistent benefits for these co-occurring disorders^1^. In contrast, behavioral treatments, primarily cognitive behavioral therapy for insomnia (CBT-I), have consistently shown efficacy in this population. CBT-I, the recommended first-line treatment for insomnia^2,3^, consists of behavioral and cognitive strategies targeting factors that perpetuate insomnia over time^4^.

Four randomized-controlled trials (RCTs) have demonstrated that CBT-I significantly improves insomnia in AUD following treatment^5–8^. Two of these trials showed that the benefits persisted for 6 months after the treatment concluded^5,7^. Although none of these trials reported a change in drinking outcomes, a recent study showed improvement in drinking-related problems after treatment^8^. Despite the promising finding, all these behavioral studies had significant methodological limitations that limited the internal validity of CBT-I treatment in individuals with AUD. The limitations included unreported or a minimal number of underrepresented participants^5,8^; subjects with past-month polysubstance use such as cocaine, amphetamine and opioids^8^; samples of 11 or fewer subjects per study arm^6,7^; a heterogeneous population that varied from early to sustained remission from alcohol use^5,6^; a failure to screen for other common sleep disorders, such as sleep-related breathing disorders^8^; inclusion of subjects from controlled environments with regimented sleep-wake hours and limited to access and cues for alcohol consumption^8^; required sobriety from alcohol for ≥1 month^5^; a study design with unequal duration of therapist follow up between treatment arms^5,8^; and a failure to account for medical comorbidity, which is prevalent in this population^5–8^.

Individuals with AUD are most vulnerable to relapse during the first few months of recovery^9,10^, a period commonly characterized by insomnia and increased urge to drink (alcohol craving)^11,12^. In recent studies of actively drinking, treatment-seeking patients with AUD, insomnia was positively associated with craving^13,14^. However, it remains unclear whether improving insomnia through behavioral treatment can reduce the intensity of craving, or other drinking-related outcomes.

We conducted a prospective RCT of standard CBT-I in Veterans with AUD during the first year of recovery. We used a randomized, 8-week intervention (CBT-I vs. Quasi-Desensitization Therapy (QDT)), with post-treatment follow-up visits at 3 and 6 months. Treatment was provided by a senior behavioral sleep medicine provider with extensive experience administering behavioral sleep treatment in patients with substance use disorders. The study aims were to: 1) Determine the efficacy of an 8-week CBT-I treatment for improving insomnia and its durability at 3- and 6-month post-treatment follow-up. 2) Investigate whether CBT-I treatment enhanced alcohol-related outcomes. 3) Evaluate, on a secondary basis, whether treatment with CBT-I improved measures of daytime functioning such as self-reported well-being, depressive symptoms, and anxiety symptoms. We also explored the role of insomnia treatment outcome on alcohol abstinence and craving.

## Methods

### Institution

The study was conducted at the Cpl. Michael J Crescenz VA Medical Center (CMCVAMC) in Philadelphia, PA between January 2015 – August 2020. The Institutional Review Board at CMCVAMC approved the conduct of the study, and all subjects gave informed consent before participating in the study. The study was registered on ClinicalTrials.gov: NCT01987089.

### Design

This 8-week, randomized, single-blind, parallel-group, outpatient trial of CBT-I or QDT involved individuals seeking treatment for AUD with insomnia. On successful completion of screening over 3-4 weeks, subjects were randomly assigned to receive CBT-I (active treatment) or QDT (comparator treatment) in 8 weekly in-person sessions. They returned for post-treatment follow-up visits 3 and 6 months later.

### Randomization and Treatment Allocation

Patients were block randomized 1:1 to CBT-I or QDT. The study statistician (KHM) created a randomization sequence using a pseudo-random number generator with block sizes of 4. Treatments were sequentially allocated in sealed envelopes by a staff member not involved in study-related assessments. All staff members who evaluated patients were blinded to treatment assignment.

### Subjects

Subjects were Veterans with moderate to severe AUD, 21-70 years of age, able to give informed consent, with a self-reported sleep latency or wake time after sleep onset >30 min on three or more nights per week for ≥1 month, an Insomnia Severity Index (ISI)^15^ total score of ≥15 (moderate to severe insomnia), abstinent from heavy drinking for ≥4 weeks and overall abstinence <1 year, and not in acute alcohol withdrawal (Clinical Institute Withdrawal Assessment scale for alcohol-revised total score^16^ <8, at baseline). Most were receiving psychosocial treatment for drinking in the outpatient clinic. Subjects were excluded from the study if they met past-year criteria for any other moderate to severe substance use disorder (tobacco use or episodic cannabis use was permissible); had a urine drug screen positive for drugs of abuse; met lifetime diagnostic criteria for Bipolar I or II disorder, Schizophrenia, or another psychotic disorder, as determined on the SCID-I^17^; had an unstable medical diagnosis; were taking medications known to affect drinking outcomes^18^, had evidence of severe cognitive impairment as assessed by the Blessed Orientation-Memory-Concentration test weighted score ≥16; or were untreated or treatment-nonadherent diagnosed with moderate to severe obstructive sleep apnea (OSA).

### Interventions

A) <u>The CBT-I treatment</u> was based on the CBT-I treatment manual used at the Penn Behavioral Sleep Medicine clinic^4^. A certified, senior behavioral sleep medicine provider provided individual CBT-I treatment to all subjects. Session 1 was an orientation session in which Sleep Restriction Therapy (SRT)^19^ and Stimulus Control therapy^20^ were initiated. Session 2 covered Sleep Hygiene^21^. Sessions 3 and 4 delivered a specific form of *cognitive therapy*^4,22^. Sessions 4-7 ensured patient adherence and titration of the optimal time in bed. The final session focused on relapse prevention strategies for insomnia. B) <u>Quasi-Desensitization Therapy (QDT)</u>. This form of therapy has been commonly used as a comparator in prior studies of behavioral interventions for insomnia^23^. During session 1 the therapist presented the QDT to eliminate “conditioned arousal,” occurring after nocturnal arousal using 8 sessions, administered weekly like the CBT-I sessions. In session 1, the therapist helped the subject develop a chronological 12-item hierarchy of commonly practiced activities on awakening at night. As a next step, the subject developed 6 imaginal scenes of engaging in neutral activities, like reading a book. During sessions 2-8, the therapist helped the subject pair these neutral scenes with the items from the 12-item hierarchy, which the subject practiced ≥2 hours before bedtime.

### Assessments

*A) **<u>Screening phase.</u>*** 1) <u>Structured Clinical Interview for DSM-IV-TR Axis I Disorders (SCID-I)</u>^17^ was used to screen for past-year alcohol dependence and other substance use disorders and to assess for a lifetime diagnosis of mania or psychotic disorder. 2) <u>PTSD Check List – Specific (PCL-S)</u>^24^ was used with a cut-off score of ≥ 40 to diagnose PTSD. 3) <u>Home Sleep Testing.</u> A type 3 portable monitor was used to screen subjects for moderate-to-severe obstructive sleep apnea^25^. 4) <u>Charlson’s Comorbidity Index</u>^26^ evaluated the subjects’ medical comorbidities. *B) **<u>Treatment and Post-treatment Follow-up phase.</u>*** i) *<u>Insomnia.</u>* 1) <u>Insomnia Severity Index (ISI)</u>^15^. This 7-item, self-report questionnaire yields a total score of 0-28, with higher scores representing greater severity. The ISI total score was the primary outcome measure. The ISI was recorded during screening, every 2 weeks during treatment, and at both post-treatment follow-up visits. 2) <u>Sleep Diary.</u> Subjects completed the Perelman School of Medicine version of the daily sleep diary, which yielded the following study variables: TTB (Time To Bed), TOB (Time Out of Bed), Sleep Onset Latency (SOL, time in min required to initially fall asleep),

Wake After Sleep Onset time (WASO, the duration of wakefulness during the night, recorded in min), Total Sleep Time (TST, total time of sleep each night, in min), and Sleep Efficiency (SE, TST/TIB). Time in Bed (TIB) was calculated as the difference between TOB and TTB. The sleep diary variables used in this study included SOL, WASO, TST and SE. The one-week sleep diary was collected at screening, every treatment visit and at the 3- and 6-month post-treatment follow-up visits. ii) *<u>Alcohol Use.</u>* 1) <u>Time-Line Follow Back interview (TLFB)</u>^27^. The TLFB uses a calendar format to estimate the number of standard drinks consumed per day. In screening, we assessed the 90 drinking days during the past year in the screening phase, the number of days abstinent before commencing sleep treatment, weekly drinking during the 8-week treatment phase and for 90 days at the 3- and 6-month follow-up visits. Variables extracted from the TLFB were the percent days abstinent (PDA, primary outcome measure), drinks per day, and percent non-heavy drinking days (PNHDD). Heavy drinking was defined as the consumption of ≥ 5 drinks in a day for men and ≥ 4 drinks per day for women. 2) <u>Penn Alcohol Craving Scale (PACS)</u>^28^. This 5-item, self-report measure was used to evaluate alcohol craving over the prior 7 days at all assessment visits. 3) <u>Short Index of Problems (SIP)</u>^29^. This 15-item, self-report instrument evaluated alcohol-related problems over the last 12 weeks, at screening and the week-8 post-treatment visit. iii) *<u>Daytime Functioning.</u>* 1) <u>12-item Short Form survey (SF-12)</u>^30^. This self-report instrument queried physical and mental wellbeing with the Physical and Mental Composite Scores (PCS and MCS), at screening, weeks 4 and 8 of treatment, and the 3- and 6-month post-treatment follow-up visits (primary outcome measure of daytime functioning). 2) <u>Beck Depression Inventory (BDI – II)</u>^6,31^. This 21-item self-report questionnaire assessed daytime mood, yielding a total score of 0-63, with higher scores indicative of more severe mood disturbance. We administered it at screening and all subsequent study-related visits and used the total score without the sleep item in our analysis. 3) <u>Trait subscale from the State-Trait Anxiety Inventory (STAI)</u>^32^. This 20-item, subject-rated scale was used to evaluate anxiety symptoms at screening, and at all treatment and post-treatment visits.^33^ C) ***<u>Treatment Fidelity, Credibility and Patient Expectation.</u>*** 1) <u>Treatment Fidelity.</u> Subjects completed a 15-item true/false sleep knowledge questionnaire at sessions 1 and 8 that was designed and used previously by the co-author (JTA)^6^. It queries sleep and insomnia features and the appropriateness of sleep-related habits addressed in the CBT-I condition. 2) <u>Treatment Credibility.</u> Subjects completed the 7-item Therapist Evaluation Questionnaire (TEQ)^34^. The first five items of the TEQ assess comprehension of the logic of the treatment, confidence in its success, resoluteness in recommending it to a friend, willingness to undergo treatment, and perceived success at treating others’ insomnia. The last two questions evaluate the therapist’s warmth/caring and confidence in the therapist’s skills. Items are rated from 1-7, with higher scores indicative of more positive feelings. Subjects completed the first five questions at treatment session 1 and all seven items at session 8. 3) <u>Patients’ Expectation.</u> The patient’s expectation were assessed in the treatment groups prior to treatment initiation (Session 1) using a modified version of the Credibility/Expectancy Questionnaire (EQ)^6,35^.

### Adverse Events

We documented all adverse events that required medical attention during the trial.

### Adherence

1) <u>Therapist Adherence to Protocol.</u> All sessions were videotaped with the subjects’ consent. The two treatment arms were divided into three blocks of 10 subjects each. The P.I. randomly selected one subject from each block (i.e., three subjects each from the CBT-I and QDT arms) to review all treatment sessions for adherence with a checklist of the elements of the manualized treatment protocols and, for content overlap between the CBT-I and QDT treatments. Our *a priori* expectation was that the sessions would cover ≥90% of the content covered by the treatment checklist, with no overlap between the CBT-I and QDT treatments. 2) <u>Patient adherence to CBT-I Treatment.</u> Adherence to sleep restriction therapy within CBT-I was assessed by computing the difference between the prescribed Time-in-Bed and the reported Time-in-Bed on the sleep diaries. Deviations ≥105 min each week were considered noncompliant for that week, consistent with prior studies^7,36^. Similarly, on an exploratory basis, adherence to Stimulus Control therapy was calculated as the difference between “wake after sleep onset” and “time spent out of bed during the night” for each night. Deviations ≥105 min each week were considered noncompliant for that week.

### Statistical Analyses

<u>Power calculation</u>: An *a priori* power calculation based on a prior study^6^ estimated that 60 subjects (30 per group) provided power adequate to detect clinically relevant differences between the treatment groups for Aims 1-3. This calculation assumed 80% power, a 2-sided p-value of 0.05, and a detectable difference (standard deviation) of the change of 1.90 (3.80) points on the ISI (Aim 1), 7.98 (15.96) % for PDA (Aim 2), and 4.48 (9.15) points for health-related quality of life (Aim 3). <u>Missingness</u>: We used logistic regression models with missingness as the binary outcome and outcome measures as individual predictors. Occasional missing values were imputed using a threshold of >95% available data and the mean of the preceding and following responses. <u>Analysis of treatment:</u> We used the intent-to-treat principle to compare treatment groups and included all randomized subjects. Baseline variables imbalanced across groups were treated as confounders and included in the models. The two primary outcome variables were ISI total score (insomnia), and PDA (alcohol use). The secondary outcomes were: 1) sleep-related (SOL, WASO, TST, and SE), 2) alcohol-related (PNHDD and PACS total score), and 3) daytime functioning (PCS, MCS, BDI total score, and STAI-trait scale) variables. Linear mixed-effects regression models employing maximum likelihood estimation or generalized estimating equations (GEE) with robust error estimation were used, as appropriate, with 999-iteration boot-strapped confidence intervals to compare the outcome trajectory between treatment groups across up to 7-time points (baseline, 4 treatment weeks, and 3- and 6-month post-treatment follow-up visits) for ISI and 10 time points (eight treatment weeks, and 3- and 6-month post-treatment periods) for the remaining measures. Each model evaluated categorical, continuous, quadratic, and spline time trends and treatment-by-time interactions as a formal test of the intervention effect, using an unstructured variance-covariance matrix structure of random effects for subject and time, and the Bayesian Information Criterion for the final model selection. We present the results as the difference between the model-estimated change from the baseline and their 95% confidence intervals. Given their Gamma distribution, WASO and PDA were modeled over time and across treatment groups using GEEs with a natural log-link function and an independent within-group correlation structure and using 10 timepoints for WASO (eight treatment weeks, and 3- and 6-month post-treatment periods), and four time-intervals for PDA (90 days in the pretreatment phase, 56 days in treatment, and 90 days each for 3- and 6-month post-treatment periods). We used the predicted value for each outcome from the longitudinal analysis to compute difference scores. The within-group effect sizes were calculated using the mean and standard deviation of change in their observed values for week 8 and 3- and 6-month follow-up visits, relative to their baseline values, with values of 0.20, 0.50, and 0.80 representing the cutoff for small, moderate, and large effect size estimates^7,37^. <u>Effect of Insomnia Treatment on Alcohol Outcomes.</u> Remission from insomnia was defined as a model-estimated subject-specific ISI total score <8, while treatment response was defined as a decrease of >7 points in the model-predicted change in ISI total score relative to pretreatment scores^38^. We divided the sample into three mutually exclusive subgroups based on treatment response at week 8, i.e., remitters (N = 24), responders (N = 12), and non-responders (N = 20), and excluded data from insomnia treatment outcome from the seven subjects with missing insomnia or drinking data at or beyond week 8. GEE models evaluated PDA (primary) and PACS total score (secondary) outcomes across the three groups over the three post-treatment time points and the associated group x time interactions using days abstinent before the onset of treatment and baseline WASO as covariates. <u>Mediation analysis.</u> Two exploratory mediation analyses were proposed to evaluate: 1) Whether a change in ISI mediated the relationship between treatment groups and SIP total score after treatment, as shown previously^8^ and 2) Whether a change in sleep diary variables (SOL, WASO, or TST) mediated the relationship between improvements in the ISI total score and alcohol outcomes after treatment (PDA, PACS total score). We used Stata 17.0 for statistical analyses.

## Results

### Subjects

We randomly assigned 63 subjects to the CBT-I (N= 31) or QDT (N=32) conditions (Supplementary Figure S1). Fifty-nine subjects (94%) completed the 8-week treatment period, and 55 subjects (87%) completed the 3-month and 57 (90%) completed the 6-month post-treatment follow-up visits. Seven subjects dropped out of the study, mostly from the QDT arm (6/7, p = 0.05). Dropouts did not differ significantly from completers on age, race, marital status, baseline ISI total score, WASO, or PDA. On average, subjects were middle-aged, male, and African American (reflecting the racial demographic of Philadelphia), whose insomnia was of moderate severity (Table 1). When compared to the QDT group, the CBT-I group had greater baseline WASO (44.58±30.02 Vs 28.60±25.51 minutes, p = 0.02) and shorter abstinence (86.67±103.32 Vs 142.68±111.64 days, p = 0.04). There was occasional missing data and no association between missingness and outcomes measures for each aim.

**Table 1.** Baseline Demographics and Clinical Characteristics (N=63)

| Characteristics | Total Sample | Treatment Groups |  | p-value |
| --- | --- | --- | --- | --- |
| **Demographic** | (n= 63) | QDT (n=32) | CBT-I (n=31) |  |
| Age (years) | 51.60 (9.59) | 51.56 (8.27) | 51.64 (10.93) | 0.97 |
| Gender (male), N (%) | 58 (92.06%) | 30 (47.62%) | 28 (44.44%) | 0.67 |
| Race (African American), N (%) | 52 (82.54%) | 29 (46.03%) | 23 (36.51%) | 0.10 |
| Ethnicity (Hispanic) | 2 (3.17) | 1 (1.59) | 1 (1.59) | 0.74 |
| Service branch (Persian Gulf and younger) | 26 (41.27%) | 15 (23.81%) | 11 (17.46%) | 0.52 |
| Marital Status (single), N (%) | 54 (85.71%) | 25 (39.68%) | 29 (46.03%) | 0.14 |
| Education (years), N (%) | 12.89 (1.44) | 13.22 (1.92) | 12.60 (0.84) | 0.38 |
| Employment Status (unemployed), N (%) | 27 (52.94%) | 11 (21.57%) | 16 (31.37%) | 0.21 |
| **Sleep** |  |  |  |  |
| Insomnia Severity Index total score | 19.38 (3.63) | 19.06 (3.77) | 19.70 (3.52) | 0.48 |
| *Sleep diary measures* |  |  |  |  |
| Sleep Onset Latency (SOL) | 50.47 (37.69) | 49.33 (34.09) | 51.58 (41.41) | 0.81 |
| Wake After Sleep Onset time (WASO) | 36.72 (28.81) | 28.60 (25.51) | 44.58 (30.02) | **0.02** |
| Total Sleep Time (TST) | 330.81 (95.46) | 345.13 (99.52) | 316.96 (90.82) | 0.25 |
| Sleep Efficiency (SE) (N=61) | 70.59 (15.16) | 72.20 (14.92) | 69.03 (15.46) | 0.41 |
| **Alcohol Consumption** |  |  |  |  |
| Days abstinent before the onset of treatment | 115.22 (110.40) | 142.68 (111.64) | 86.87 (103.32) | **0.04** |
| Percent Days Abstinent | 32.52 (32.98) | 26.90 (29.20) | 38.31 (36.03) | 0.17 |
| Penn Alcohol Craving Scale total score (n=61) | 9.50 (7.34) | 8.22 (7.03) | 10.83 (7.53) | 0.16 |
| Drinks per Day | 8.99 (8.17) | 8.82 (7.10) | 9.18 (9.26) | 0.86 |
| Percent Heavy Drinking Days | 54.85 (39.32) | 56.45 (39.84) | 53.18 (39.37) | 0.74 |
| Short Index of Problems total score (n = 56) | 19.53 (14.21) | 20.07 (14.96) | 19.03 (13.71) | 0.78 |
| **Daytime Dysfunction & Psych. Symptoms** |  |  |  |  |
| Physical Composite Score, SF-12 scale (N=39) | 41.72 (5.03) | 40.40 (5.37) | 42.75 (4.62) | 0.15 |
| Mental Composite Score, SF-12 scale (N=39) | 43.91 (6.99) | 43.18 (6.40) | 44.47 (7.51) | 0.56 |
| Beck Depression Inventory total score (N=62) | 19.04 (11.83) | 17.74 (10.80) | 20.35 (12.82) | 0.38 |
| Trait subscale (State-Trait Anxiety Inventory)<sup>1</sup> | 45.68 (5.94) | 46.30 (6.01) | 45.09 (5.92) | 0.43 |
| PCL-5 total score (without sleep items)<sup>1</sup> | 41.19 (14.42) | 40.40 (16.01) | 42.00 (12.78) | 0.66 |
| PTSD diagnosis N (%) | 31 (49.21%) | 16 (25.40%) | 15 (23.81%) | 0.89 |
| **Medical Comorbidities** |  |  |  |  |
| Charlson's Comorbidity Index total score | 1.92 (1.71) | 1.96 (1.67) | 1.87 (1.78) | 0.82 |
| **Hypnotic Medication Prescriptions (N=63)** |  |  |  |  |
| Any sedating medication, N (%) | 39 (39.00%) | 19 (19.80%) | 20 (19.20%) | 0.67 |
| Gabapentin, N (%) | 22 (34.92%) | 12 (19.05%) | 10 (15.87%) | 0.66 |
| Mirtazapine, N (%) | 8 (12.70%) | 3 (4.76%) | 5 (7.94%) | 0.47 |
| Trazodone, N (%) | 11 (17.46%) | 4 (6.35%) | 7 (11.11%) | 0.33 |
| Prazosin, N (%) | 6 (9.52%) | 2 (3.17%) | 4 (6.35%) | 0.42 |
| Hydroxyzine, N (%) | 3 (4.76%) | 2 (3.17%) | 1 (1.59%) | 1.00 |
| Melatonin, N (%) | 3 (4.76%) | 2 (3.17%) | 1 (1.59%) | 1.00 |
| Diphenhydramine, N (%) | 2 (3.17%) | 0 (0.00%) | 2 (3.17%) | 0.23 |
| Amitriptyline, N (%) | 1 (1.59%) | 1 (1.59%) | 0 (0.00%) | 1.00 |
| Quetiapine, N (%) | 1 (1.59%) | 0 (0.00%) | 1 (1.59%) | 0.49 |

Footnote: <sup>1</sup> = total score of the individual scales after excluding the sleep items;

### Sleep Outcomes

1) <u>Insomnia Severity Index.</u> The ISI total score decreased over time in both treatment groups, with no significant group by time interaction (Table 2 and Figure 1A). The within-group effect sizes at the end of treatment, and 3-month and 6-month follow-up visits for the CBT-I group were -1.63, -1.17 and -0.94 compared to -1.50, -1.64, and -1.24 for the QDT group (Table 3). The proportion of subjects who remitted from insomnia at the end of treatment was 48% in the CBT-I group, and 40% in the QDT group (p = NS). 2) <u>Sleep diary variables.</u> Similarly, SL, WASO, TST and SE improved in both treatment groups but without any treatment group by time interaction. WASO showed a greater improvement in the CBT-I treated group that was significant only at the 6-month post-treatment follow-up visit (Table 2 and Figure 1B).

**Figure 1.**
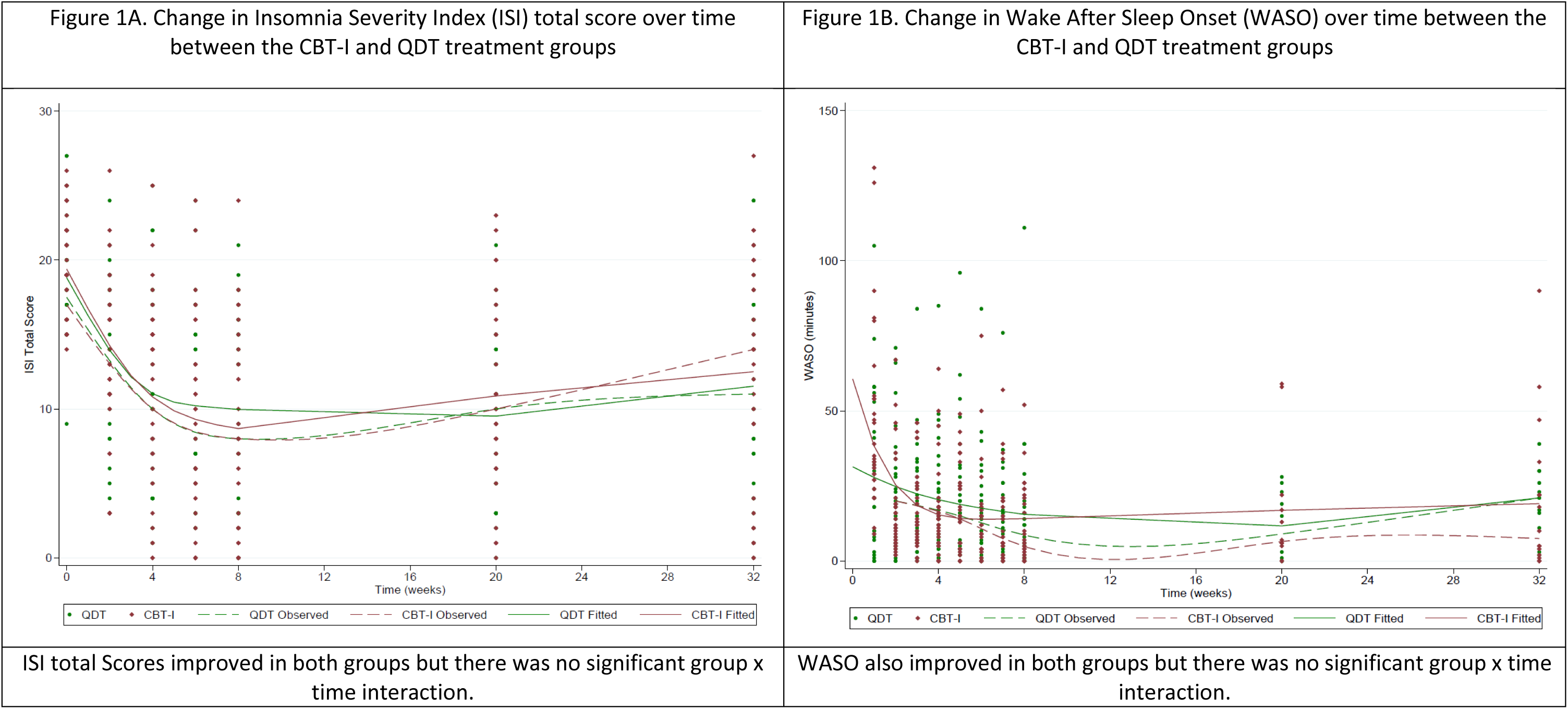
Change in sleep indices over time between the CBT-I and QDT treatment groups

**Table 2.** The difference in the change scores of variables between treatment groups at end of treatment, 3- and 6-month post-treatment follow-up visits. CBT-I = Cognitive Behavioral Therapy for Insomnia; QDT = Quasi-Desensitization Therapy; ISI = Insomnia Severity Index; SOL = Sleep Onset Latency; WASO = Wake After Sleep Onset time; TST = Total Sleep Time; SE = Sleep Efficiency; PDA = Percent Days Abstinent; PHDD – Percent Heavy Drinking Days; PACS = Penn Alcohol Craving Scale total score; PCS = Physical Composite Summary Score (from the Short Form 12-item measure); MCS = Mental Composite Summary Score (from the Short Form 12-item measure); BDI = Beck Depression Inventory – II; BDI Tot. Score = Beck Depression Inventory total score excluding the sleep item; STAI-trait = State Trait Anxiety Inventory – Trait subscale; GEE (gamma) = Generalized Estimating Equation-gamma family models; GEE (normal) = Generalized Estimating Equation-normal family models; Mixed – Linear Mixed Effects Regression models. Given the imbalance in the two variables between the treatment arms before treatment, all the models were adjusted for the number of days abstinent at screening and baseline WASO, except for the model for WASO which was only adjusted for number of days abstinent at screening. In all cases, the p-value for the time by treatment interaction was not statistically significant.

| Outcome | Model | Difference in change from baseline between CBT-I and QDT at 8 weeks |  | Difference in change from baseline between CBT-I and QDT at 3 months |  | Difference in change from baseline between CBT-I and QDT at 6 months |  |
| --- | --- | --- | --- | --- | --- | --- | --- |
|  |  | Model-based estimate | 95% Confidence Interval | Model-based estimate | 95% Confidence Interval | Model-based estimate | 95% Confidence Interval |
| ISI total score | Mixed | -1.18 | -4.11, 1.90 | 0.09 | -3.26, 3.62 | 0.45 | -3.22, 3.97 |
| SOL | GEE (gamma) | -24.36 | -72.43, 21.56 | -21.43 | -75.01, 30.92 | -27.05 | -78.29, 18.97 |
| WASO | GEE (gamma) | -40.50 | -92.61, 3.83 | -36.23 | -90.27, 7.94 | -41.22 | -91.46, -2.40 |
| TST | Mixed | -17.60 | -67.47, 40.95 | -9.38 | -96.90, 65.94 | 40.39 | -105.49, 102.68 |
| SE | Mixed | 0.09 | -0.06, 0.23 | 0.03 | -0.18, 0.19 | 0.03 | -0.28, 0.16 |
| PDA | GEE (gamma) | -0.12 | -0.28, 0.06 | -0.11 | -0.28, 0.07 | -0.15 | -0.32, 0.02 |
| PACS total score | GEE (gamma) | -1.77 | -4.91, 1.21 | -1.69 | -5.37, 2.51 | -1.21 | -4.62, 2.04 |
| Drinks per Day | GEE (gamma) | 0.82 | -5.41, 14.30 | 0.97 | -5.80, 13.94 | 1.27 | -5.02, 15.37 |
| Percent non-HDD | GEE (gamma) | -0.03 | -0.22, 0.16 | -0.02 | -0.21, 0.18 | -0.05 | -0.25, 0.14 |
| PCS (SF-12) | GEE (normal) | -0.03 | -0.76, 0.67 | -0.08 | -1.89, 1.67 | -0.13 | -3.03, 2.66 |
| MCS (SF-12) | GEE (normal) | -0.77 | -1.93, 0.31 | -1.93 | -4.83, 0.77 | -3.10 | -7.73, 1.24 |
| BDI total score | GEE (gamma) | 0.52 | -3.47, 4.56 | 0.11 | -3.51, 4.30 | 4.02 | -1.57, 9.01 |
| STAI-trait subscale | Mixed | 0.26 | -0.30, 0.74 | 0.65 | -0.74, 1.84 | 1.04 | -1.18, 2.95 |

**Table 3.** Within-group Effect Size Estimates for Variables in CBT-I and QDT After Treatment and at 3- and 6-month Follow-up Visits. Effect size is computed as a within-group difference relative to pretreatment values; CBT-I = Cognitive Behavioral Therapy for Insomnia; QDT = Quasi-Desensitization Therapy; ISI = Insomnia Severity Index; SOL = Sleep Onset Latency; WASO = Wake After Sleep Onset time; TST = Total Sleep Time; SE = Sleep Efficiency; PDA = Percent Days Abstinent; Percent non-HDD = Percent non Heavy Drinking Days; PACS = Penn Alcohol Craving Scale; PCS = Physical Composite Summary Score (from the Short Form 12-item measure); MCS = Mental Composite Summary Score (from the Short Form 12-item measure); BDI Total Score = Beck Depression Inventory - II total score after excluding the sleep item; STAI-trait = State Trait Anxiety Inventory – Trait subscale.

| Variable | End of treatment |  | 3-month Post-treatment |  | 6-month Post-treatment |  |
| --- | --- | --- | --- | --- | --- | --- |
|  | QDT | CBT-I | QDT | CBT-I | QDT | CBT-I |
| ISI total score | - 1.50 | - 1.63 | - 1.64 | - 1.17 | - 1.24 | - 0.94 |
| SOL | - 0.75 | - 0.97 | - 0.78 | - 0.39 | - 0.66 | - 0.70 |
| WASO | - 0.23 | - 0.88 | - 0.74 | - 0.87 | - 0.41 | - 1.39 |
| TST | 0.57 | 0.56 | 0.40 | 0.53 | - 0.005 | 0.58 |
| SE | 0.91 | 1.37 | 0.97 | 0.67 | 0.54 | 0.97 |
| PDA | 1.91 | 1.54 | 1.84 | 1.32 | 1.78 | 1.23 |
| Drinks per Day | 1.91 | 1.54 | 1.84 | 1.32 | 1.78 | 1.23 |
| Percent non-HDD | 1.34 | 1.34 | 1.19 | 1.09 | 1.21 | 1.07 |
| PACS total score | - 0.45 | - 0.49 | - 0.50 | - 0.26 | - 0.59 | - 0.51 |
| PCS (SF-12) | - 0.17 | - 0.26 | - 0.26 | - 0.18 | - 0.41 | - 0.14 |
| MCS (SF-12) | - 0.20 | - 0.31 | 0.27 | - 0.22 | 0.10 | - 0.21 |
| BDI total score | - 0.59 | - 0.67 | - 0.70 | - 0.59 | - 0.34 | - 0.58 |
| STAI-trait subscale | - 0.21 | - 0.10 | - 0.23 | - 0.18 | - 0.08 | 0.09 |

### Alcohol-related measures

During their study participation, 29 subjects (46%) relapsed to alcohol use (15 in the CBT-I arm and 14 in the QDT arm, p=NS). 1) <u>Percent Days Abstinent.</u> PDA improved in both groups over time but with no between-group differences, and with larger effect sizes in the QDT group (Tables 2 and 3). During the 8 weeks of treatment, 43 subjects reported remaining abstinent from alcohol (19 in the CBT-I group and 24 in the QDT group, χ^2^=1.3, p=NS), of whom 29 remained abstinent at the 6-month follow-up visit (12 in the CBT-I and 17 in the QDT groups). Subjects who relapsed to drinking were abstinent for a longer duration before the onset of treatment (146.1±104.8 versus 81.3±107.9, p=0.02), but did not differ from the other participants on age, marital status, race, baseline PDA or ISI score, or treatment arm. 2) <u>Drinks per Day, Percent non-Heavy Drinking Days, and Alcohol craving (PACS)</u> showed no differential improvement between groups over time. QDT had larger effect sizes at all timepoints, except for the PACS total score at week 8 (Tables 2 and 3).

### Daytime symptoms

There was no treatment effect on the PCS or MCS (from the SF-12 measure), BDI, or STAI-trait subscale total scores. The improvement effect size was medium to large for depressive symptoms, but small for the remaining measures at all time points (Tables 2 and 3).

### Treatment credibility and fidelity

There was no difference between the treatment groups for the therapist adherence to the protocol, pre-treatment EQ score, pretreatment item # 1 score of KAQ, change in KAQ total score with treatment, pre-treatment TEQ individual items or the total score or change with treatment, Supplementary Table S1. Dropouts did not differ from completers on pretreatment EQ, KAQ or TEQ total scores.

### Adherence

1) <u>Therapist adherence to protocol.</u> The mean ± SD adherence in the CBT-I and QDT sessions were 97.6 ± 3.5% and 93.8 ± 12.4%, p = NS. There was no content overlap between the CBT-I and QDT treatments. 2) <u>Patient adherence to treatment.</u> *a. Sleep Restriction.* Twelve subjects demonstrated partial noncompliance, with 75% being noncompliant for one week, especially treatment week 2. *b. Stimulus Control.* None of the subjects met the criteria for noncompliance.

### Insomnia Treatment Response

When compared to responders and non-responders, remitters had a significantly lower BDI total score and were less likely to be receiving psychotropic medication or hypnotic medication at baseline (Supplementary Table S2).

### The association between insomnia measures and drinking outcomes

Subjects who remitted from insomnia at week 8 had lower PACS total scores at all three timepoints than responders and non-responders (Figure 2). Although a similar trend was seen with PDA, the results were not statistically significant (Supplementary Figure S2).

**Figure 2.**
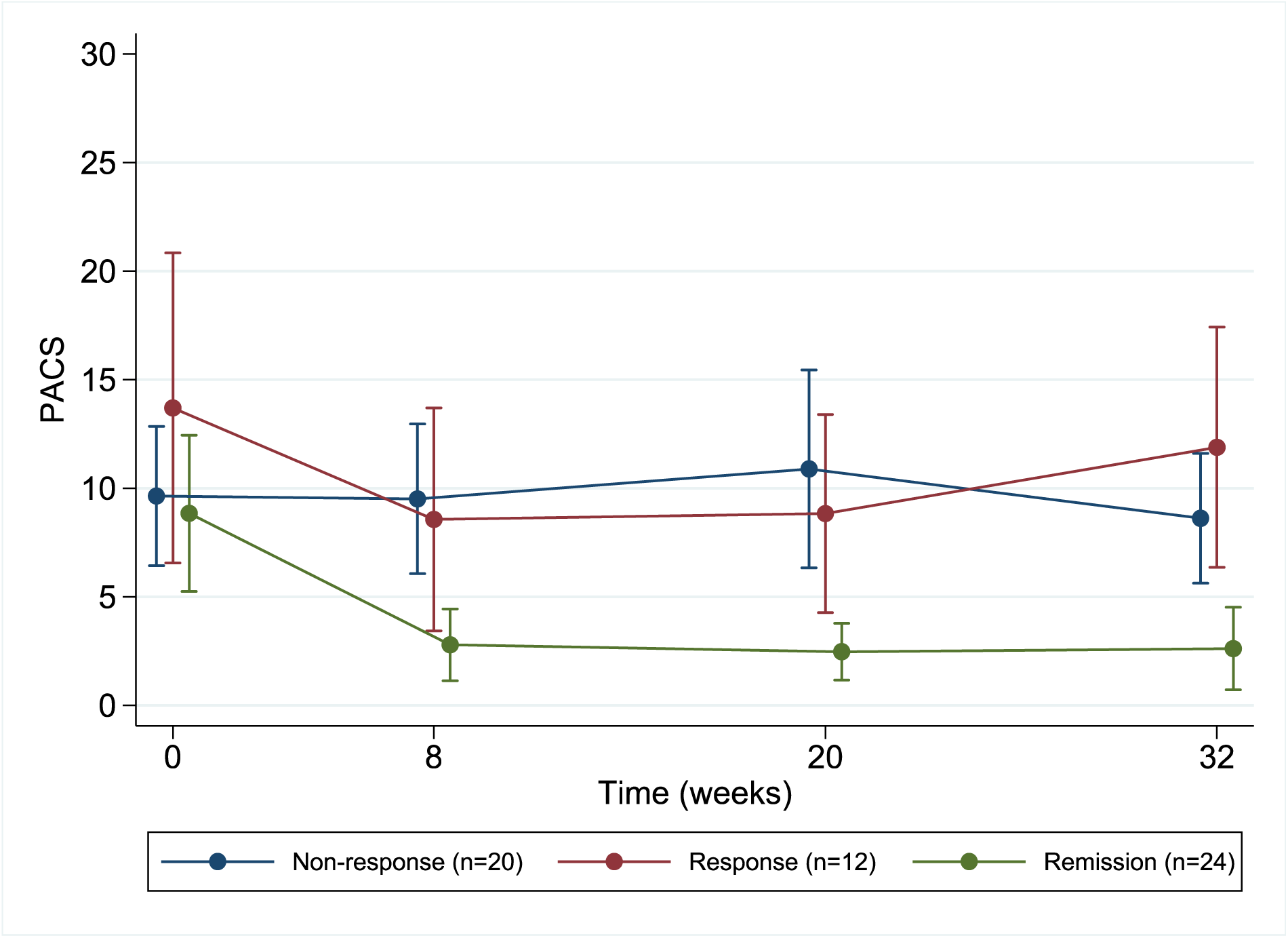
PACS total score (alcohol craving) in the groups based on insomnia treatment outcomes. *Non-response subgroup* (N = 20) included those with neither response nor remission; *Response subgroup* (N = 12) included treatments responders, i.e., those with a pre-post change in ISI<7; *remission subgroup* (N = 24) included subjects with an ISI total score <8 at week 8. Subjects with missing data who were excluded (N = 7). The results showed a statistically significant difference in PACS scores over time by group (p=0.003). When compared to the non-response group, the group that remitted from insomnia had significantly lower PACS total score at follow-up. There was no difference between the response and non-response groups.

### Exploratory mediation modeling

An improvement in the ISI total score did not mediate the relationship between treatment groups and SIP total score. Similarly, SOL, WASO and TST did not mediate the relationship between improvements in the ISI total score and alcohol-related outcomes (PDA, PACS total score).

### Adverse Events

There were no significant differences between treatment groups in reported adverse events (Supplementary Table S3).

## Discussion

This RCT evaluated the efficacy of CBT-I relative to QDT for treating insomnia, augmenting recovery from pathological drinking, and improving next-day functioning in individuals complaining of moderate to severe insomnia during early recovery from AUD. The results showed that CBT-I was associated with improvement in insomnia severity and sleep diary-related insomnia indices of magnitudes similar to those seen in prior studies. Unexpectedly, QDT also appeared to be an active treatment that was equal in efficacy to CBT-I, insofar as the treatment groups did not differ on any of the sleep indices, the number of subjects who remitted from insomnia, drinking outcomes, or daytime functioning. In the pooled sample, when compared to treatment responders and non-responders, those who remitted from insomnia had persistently decreased PACS total score during the post-treatment follow-up phase.

The current study extends prior findings from studies that used less active comparison conditions. Although CBT-I was not superior to the comparator treatment, QDT, it outperformed recent meta-analytic effect size estimates (reported as the standardized mean differences after adjusting for small sample sizes) for treatment and post-treatment follow-up periods on a variety of measures, including ISI total score, SOL, WASO, TST and SE^39,40^. Thus, the current study findings are consistent with those of pilot studies of CBT-I in treating insomnia and co-occurring AUD in veterans^7^ and non-veterans ^5,6^ in outpatient treatment settings.

QDT was initially developed as a behavioral placebo sleep treatment, but studies have shown variability in its effects on sleep. In two previous studies, CBT-I was superior to QDT on improving insomnia measures. Among subjects with insomnia, CBT-I resulted in greater improvement than QDT on reported sleep measures such as TST, middle WASO, terminal WASO, and SE^41^. In subjects with insomnia co-occurring with depression, the effect sizes of improvement in the CBT-I group were large for sleep quality, moderate for SE, and small for TST^42^. In contrast to these findings, other studies showed significant treatment effects for QDT. In subjects with insomnia and higher fatigue scale scores, QDT improved TST and SE relative to baseline scores^43^. In a pilot study of subjects with insomnia co-occurring with AUD, QDT showed large improvements on the ISI total score and a medium effect on SOL, frequency of awakenings, BDI, and STAI-trait total scores. However, it was inferior to CBT-I in improving reported SE, WASO, and daytime fatigue symptoms^6^. Here, QDT was as effective as CBT-I in improving sleep measures, with large effects at the post-treatment visit on the ISI total score, SOL, and SE. These findings suggest that, specifically in the context of insomnia in recovering AUD, QDT is not an inert intervention but an active treatment comparable in efficacy to CBT-I. More importantly, the current study extends the literature to suggest that a treatment targeting arousal, without being specifically focused on insomnia, may additionally help improve drinking outcomes. Given the heightened arousal in AUD patients and QDT’s demonstrated benefits in promoting relaxation at night and improving drinking outcomes, a reduction in arousal might be a common mechanism that alleviates both insomnia and alcohol consumption. Thus, counter-arousal strategies may serve as an important alternative treatment for insomnia in this population and offer the promise of an additional intervention for insomnia.

The lack of a difference between the treatment arms for any clinical outcome allowed us to evaluate the total sample stratified by their insomnia treatment outcomes to assess their baseline characteristics and relationship with alcohol consumption measures. The finding that a better insomnia treatment outcome was associated with greater depression severity at baseline aligns with the observation in older patients with insomnia^44^ but contrasts with the results from other studies^45,46^. It is possible that in the context of insomnia that co-occurs with AUD, a higher depressive symptom burden may represent an inability of the patient to follow through with the recommendations of behavioral sleep treatments, an observation commonly seen in the clinical setting. Further, an increased severity of depression may be associated with a greater likelihood of being prescribed psychotropic medications, as shown previously^47^. Of concern is the possibility that many psychotropic medications, e.g., serotonergic antidepressant medications, may lead to sleep disturbance by fragmenting sleep^48^. Thus, the higher rates of prescription psychotropic medications may represent an increased severity of co-occurring psychiatric disorders or sleep disturbance aggravated by iatrogenic causes that led to treatment non-response with the behavioral sleep intervention.

The early recovery phase in AUD is a critical time during which many individuals relapse to drinking. Insomnia and alcohol craving are two known risk factors for drinking that are more severe during early recovery. Prior studies have shown that insomnia is positively correlated with craving, but treatment with quetiapine has shown variable effects on this relationship^13,14^. The current study is the first to show that remission from insomnia with behavioral sleep treatment is associated with a consistently lower intensity of alcohol craving over the 6-month post-treatment period and a non-significant improvement in the percent days abstinent. This association is plausible as alcohol consumption initially triggers dopamine release in the nucleus accumbens and the ventral tegmental area (VTA). However, persistent drinking leads to decreased dopamine release, sleep disturbances, increased alcohol cravings, and an escalation in consumption to stimulate further dopamine release. The GABAergic neurons in the rostro-medial tegmental nucleus, rich in gamma-aminobutyric acid, modulate dopamine release in the VTA^5051^ and influence arousal. This modulation may become abnormal during pathological drinking but normalizes with abstinence and improved sleep continuity, thereby reducing cravings^52^.

Strengths of the current study are that it is the largest randomized, controlled trial of CBT-I for insomnia in an outpatient naturalistic treatment setting with a predominantly African-American sample, it used objective monitoring to screen out moderate-to-severe obstructive sleep apnea and an active treatment control condition, treatment fidelity was high, treatment effects were not altered by medications for drinking, subject retention was high, and the post-treatment observational phase was 6 months. Despite these strengths, the selection of treatment-seeking individuals may limit the generalizability of these findings to all individuals with AUD and insomnia although one recent study showed that CBT-I improved insomnia in non-treatment-seeking individuals with AUD or other substance use disorders^53^. The subjective nature of the outcome measures and the lack of collateral information may have introduced a reporting bias on the subjects’ sleep and alcohol-related outcomes. Incorporating objective sleep monitoring, a biological measure of alcohol consumption, such as phosphatidylethanol, and using a method to measure compliance to QDT treatment, could enhance both the procedures and the findings. Future studies with larger sample sizes and collateral information may demonstrate between-group differences in alcohol consumption and enable an exploration of the underlying mechanisms of the CBT-I and QDT treatments.

## Conclusion

We found that CBT-I and QDT are equally effective in treating insomnia in patients in early recovery from AUD. Thus, QDT, a counter-arousal strategy is also effective in this population. Subjects who achieved remission from insomnia had a lower intensity of depressive symptoms, were less likely to be on psychotropic medications at baseline and had consistent improvements in alcohol craving for up to 6 months post-treatment.

## Data Availability

Any dataset from this study cannot be released as per the directive of the IRB at CMCVAMC.

## Contributors

SC, MLP and JTA conceptualized the study; SC, SH and JF evaluated subjects; SC, KHM, JTA and MLP analyzed data; SC, KHM, JTA, MLP, HRK, DWO, STK and JF collaborated in the generation of the final manuscript.

## Acknowledgment

We thank Mr. Elliott Sturgis, Mr. Walliuddin Khader, Ms. Brittany Taylor and Ms. Nina Thakur for their assistance with the data collection. Role of funding sources: The VA Clinical Sciences Research and Development grant IK2CX000855 provided funding for the conduct of this trial (SC) and VA grant 5I01CX001957 provided time for data analysis and write up of the manuscript. Support from NIH grants R01 AG041783 (to M.L.P.), R56 AG050620 (to M.L.P.); and R01 AA023192 and R01 AA021164 (to H.R.K.) was provided to co-authors of this study.

## Disclosure Statement

None of the authors report any conflict of interest with this investigation. Dr. Chakravorty has received research support from AstraZeneca and Teva pharmaceuticals. Dr. Perlis has received research support from Nexalin technology and Teva pharmaceuticals. Dr. Kranzler is a member of advisory boards for Altimmune, Clearmind Medicine, Dicerna Pharmaceuticals, Enthion Pharmaceuticals, Lilly Pharmaceuticals, and Sophrosyne Pharmaceuticals; a consultant to Sobrera Pharmaceuticals and Altimmune; the recipient of research funding and medication supplies for an investigator-initiated study from Alkermes; a member of the American Society of Clinical Psychopharmacology’s Alcohol Clinical Trials Initiative, which was supported in the last three years by Alkermes, Dicerna, Ethypharm, Imbrium, Indivior, Kinnov, Lilly, Otsuka, and Pear; and a holder of U.S. patent 10,900,082 titled: "Genotype-guided dosing of opioid agonists," issued 26 January 2021.

---

**Supplementary Table S1.**
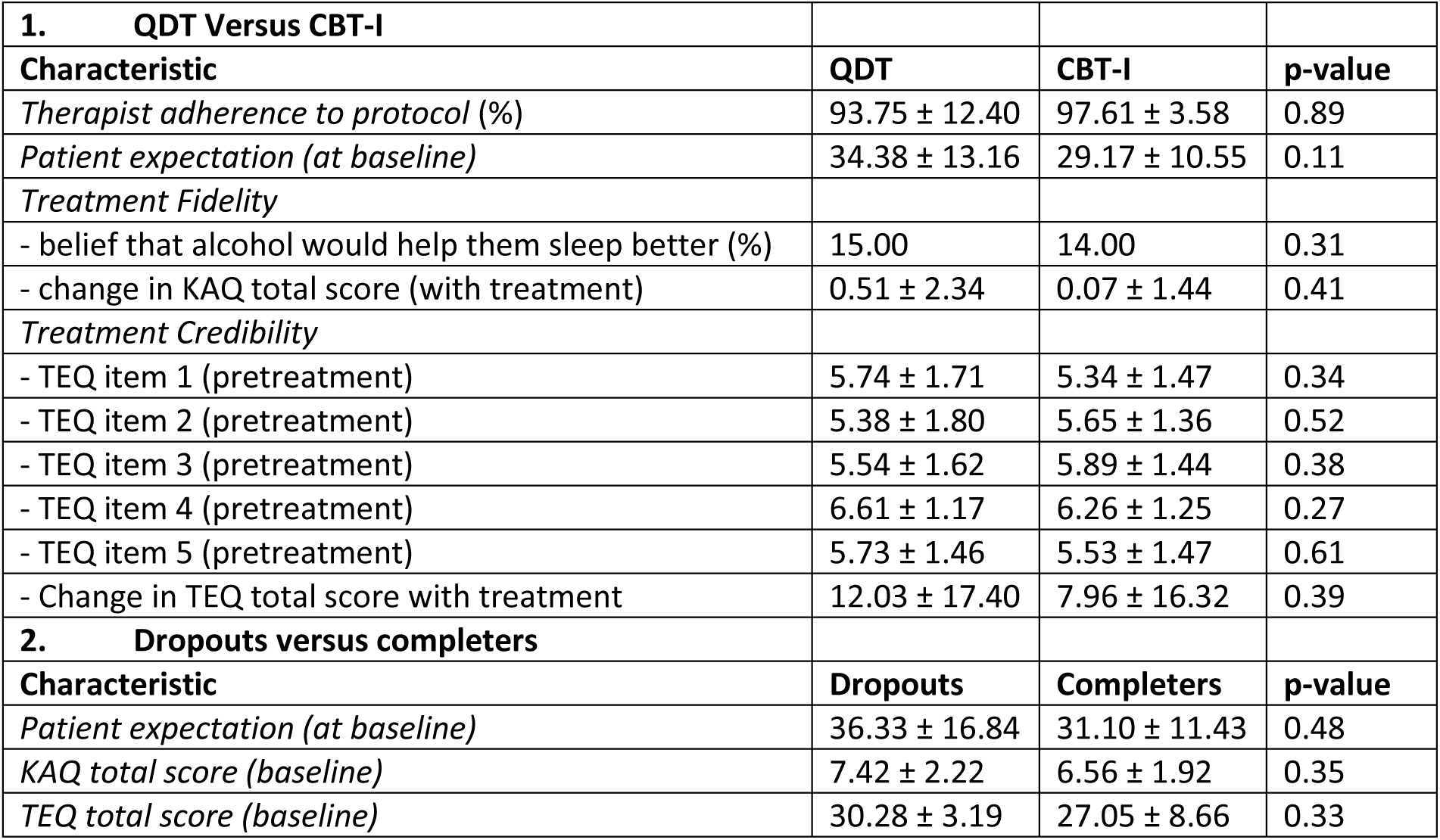
Treatment credibility and fidelity scores (mean ± standard deviation) across groups. Addendum: 1) <u>Patient adherence to treatment.</u> *a. Sleep Restriction.* Twelve subjects showed partial noncompliance, with 75% being noncompliant for one week, especially treatment week 2. *b. Stimulus Control.* No subjects met criteria for noncompliance. QDT = quasi-desensitization therapy; CBT-I = cognitive behavioral therapy for insomnia.

| **1. QDT Versus CBT-I** |  |  |  |
| --- | --- | --- | --- |
| **Characteristic** | **QDT** | **CBT-I** | **p-value** |
| *Therapist adherence to protocol (%)* | 93.75 ± 12.40 | 97.61 ± 3.58 | 0.89 |
| *Patient expectation (at baseline)* | 34.38 ± 13.16 | 29.17 ± 10.55 | 0.11 |
| *Treatment Fidelity* |  |  |  |
| - belief that alcohol would help them sleep better (%) | 15.00 | 14.00 | 0.31 |
| - change in KAQ total score (with treatment) | 0.51 ± 2.34 | 0.07 ± 1.44 | 0.41 |
| *Treatment Credibility* |  |  |  |
| - TEQ item 1 (pretreatment) | 5.74 ± 1.71 | 5.34 ± 1.47 | 0.34 |
| - TEQ item 2 (pretreatment) | 5.38 ± 1.80 | 5.65 ± 1.36 | 0.52 |
| - TEQ item 3 (pretreatment) | 5.54 ± 1.62 | 5.89 ± 1.44 | 0.38 |
| - TEQ item 4 (pretreatment) | 6.61 ± 1.17 | 6.26 ± 1.25 | 0.27 |
| - TEQ item 5 (pretreatment) | 5.73 ± 1.46 | 5.53 ± 1.47 | 0.61 |
| - Change in TEQ total score with treatment | 12.03 ± 17.40 | 7.96 ± 16.32 | 0.39 |
| **2. Dropouts versus completers** |  |  |  |
| **Characteristic** | **Dropouts** | **Completers** | **p-value** |
| *Patient expectation (at baseline)* | 36.33 ± 16.84 | 31.10 ± 11.43 | 0.48 |
| *KAQ total score (baseline)* | 7.42 ± 2.22 | 6.56 ± 1.92 | 0.35 |
| *TEQ total score (baseline)* | 30.28 ± 3.19 | 27.05 ± 8.66 | 0.33 |

**Supplementary Table S2.** Differences in baseline demographic and clinical variables across the groups based on insomnia treatment response at the end of week 8 of treatment. *Remission subgroup* (N = 24) included subjects with an ISI total score <8 at week 8; *Response subgroup* (N = 12) included treatments responders, i.e., those with a per-post change in ISI<7; *Non-response subgroup* included those with neither response nor remission; ISI = Insomnia Severity Index; SOL = Sleep Onset Latency; WASO = Wake After Sleep Onset time; TST = Total Sleep Time; Dur. Of abstinence = duration of abstinence from alcohol before onset of behavioral treatment for insomnia; PDA = Percent Days Abstinent; Percent HDD – Percent Heavy Drinking Days; PACS = Penn Alcohol Craving Scale total score; BDI total score = Beck Depression Inventory total score excluding the sleep item; STAI-trait score = State Trait Anxiety Inventory – Trait subscale score; Post-hoc testing for the BDI scores using Bonferroni’s test shows remitters < responders (p = 0.001) but remitters < non-responders (p = 0.08).

| Domain | Variable | Remission (N=24) | Response (N=12) | Non-response (N=20) | p-value |
| --- | --- | --- | --- | --- | --- |
| Demographic | Age | 52.9±10.1 | 52.7±8.9 | 51.8±8.2 | 0.76 |
|  | Gender (males) | 22 (22/24) | 10 (10/12) | 19 (19/20) | 0.60 |
|  | Marital status (single) | 20 (20/24) | 11 (11/12) | 19 (19/20) | 0.55 |
| Sleep | ISI total score | 18.5±4.2 | 21.3±3.0 | 18.8±2.8 | **0.07** |
|  | SOL | 47.3±33.2 | 61.8±55.0 | 49.0±36.7 | 0.89 |
|  | WASO | 39.0±33.1 | 28.5±23.6 | 38.3±27.7 | 0.63 |
|  | TST | 332.7±109.0 | 347.6±81.5 | 327.3±91.9 | 0.85 |
| Alcohol | Days abstinent before Tr. | 120.5±97.8 | 112.1±117.9 | 117.0±121.8 | 0.98 |
|  | PDA | 33.3±33.7 | 36.7±29.4 | 27.8±34.9 | 0.74 |
|  | Drinks per day | 9.4±8.1 | 10.3±10.3 | 8.0±7.8 | 0.66 |
|  | Percent HDD | 60.7±37.8 | 51.6±38.7 | 51.5±42.6 | 0.69 |
|  | PACS total score | 7.7±6.7 | 11.3±8.5 | 9.3±6.6 | 0.37 |
| Psychiatric | BDI total score | 12.6±9.8 | 26.1±11.2 | 19.6±10.1 | **0.001** |
|  | STAI-trait score | 46.8±4.7 | 47.3±7.2 | 43.2±6.1 | 0.08 |
|  | PTSD status (positive) | 8 (8/24) | 8 (8/12) | 11 (11/20) | 0.14 |
|  | Psychotropic meds | 9 (16.07%) | 11 (19.64%) | 15 (26.79%) | **0.002** |
| Medical | Charlson's Index | 1.8±1.5 | 2.1±1.6 | 2.2±2.0 | 0.74 |

**Supplementary Table S3.** Adverse events with treatment. CBT-I = Cognitive Behavioral Therapy for Insomnia; QDT = Quasi-Desensitization Therapy

| Event | QDT (N = 32) | CBT-I (N = 31) | p-value |
| --- | --- | --- | --- |
| Abdominal pain | 0 (0.00%) | 1 (1.59%) | 0.49 |
| Cerebrovascular accident | 1 (1.59%) | 0 (0.00 %) | 1.00 |
| Alcohol detoxification | 2 (3.17%) | 4 (6.35%) | 0.42 |
| Residential treatment for drinking | 0 (0.00%) | 1 (1.59%) | 0.49 |
| Pneumonia | 0 (0.00%) | 1 (1.59%) | 0.49 |
| Total | 3 (4.76) | 7 (11.11%) | 0.18 |

**Supplementary Figure S1.**
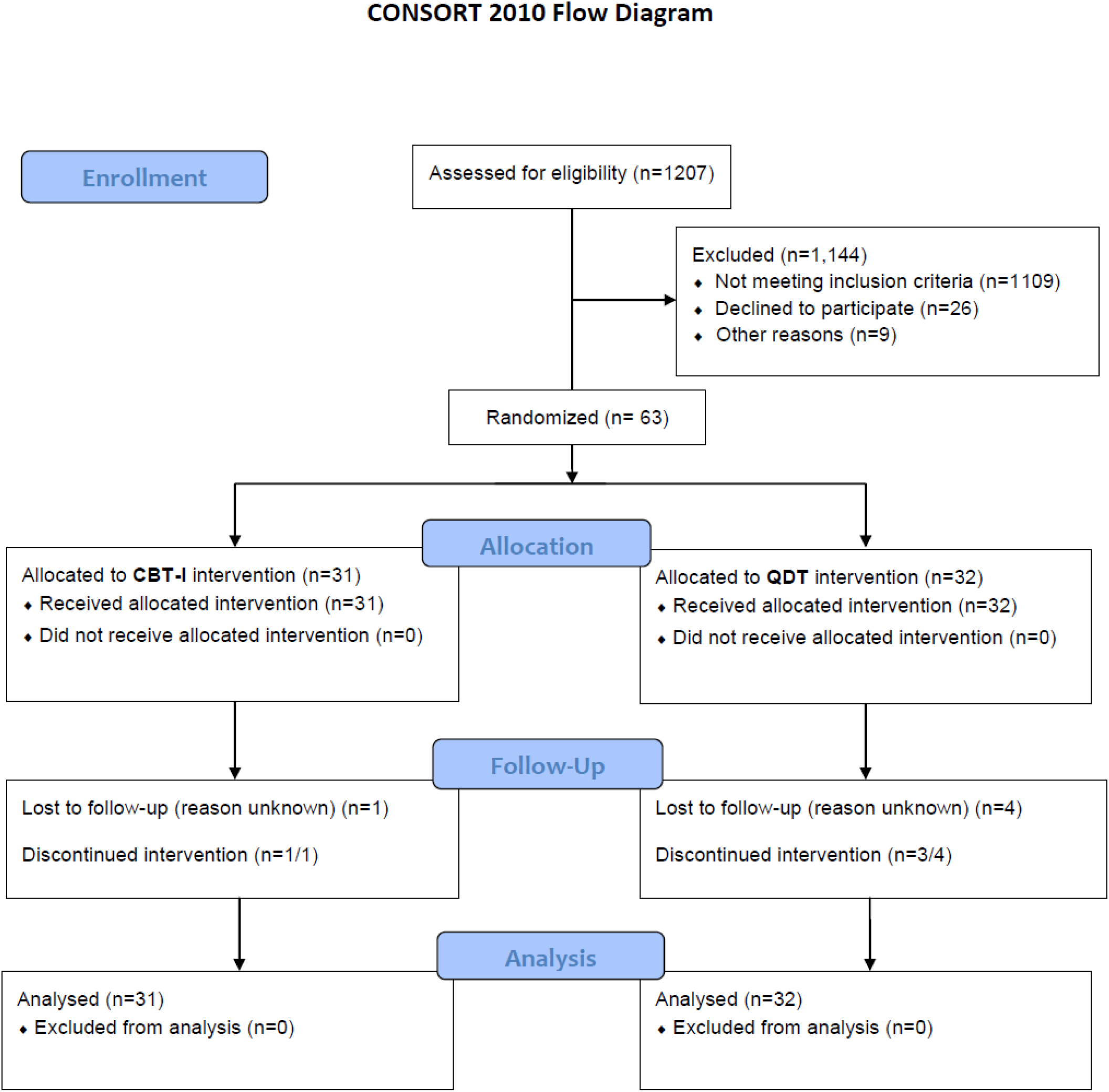
CONSORT flow chart for flow of subjects in the study

**Supplementary Figure S2.**
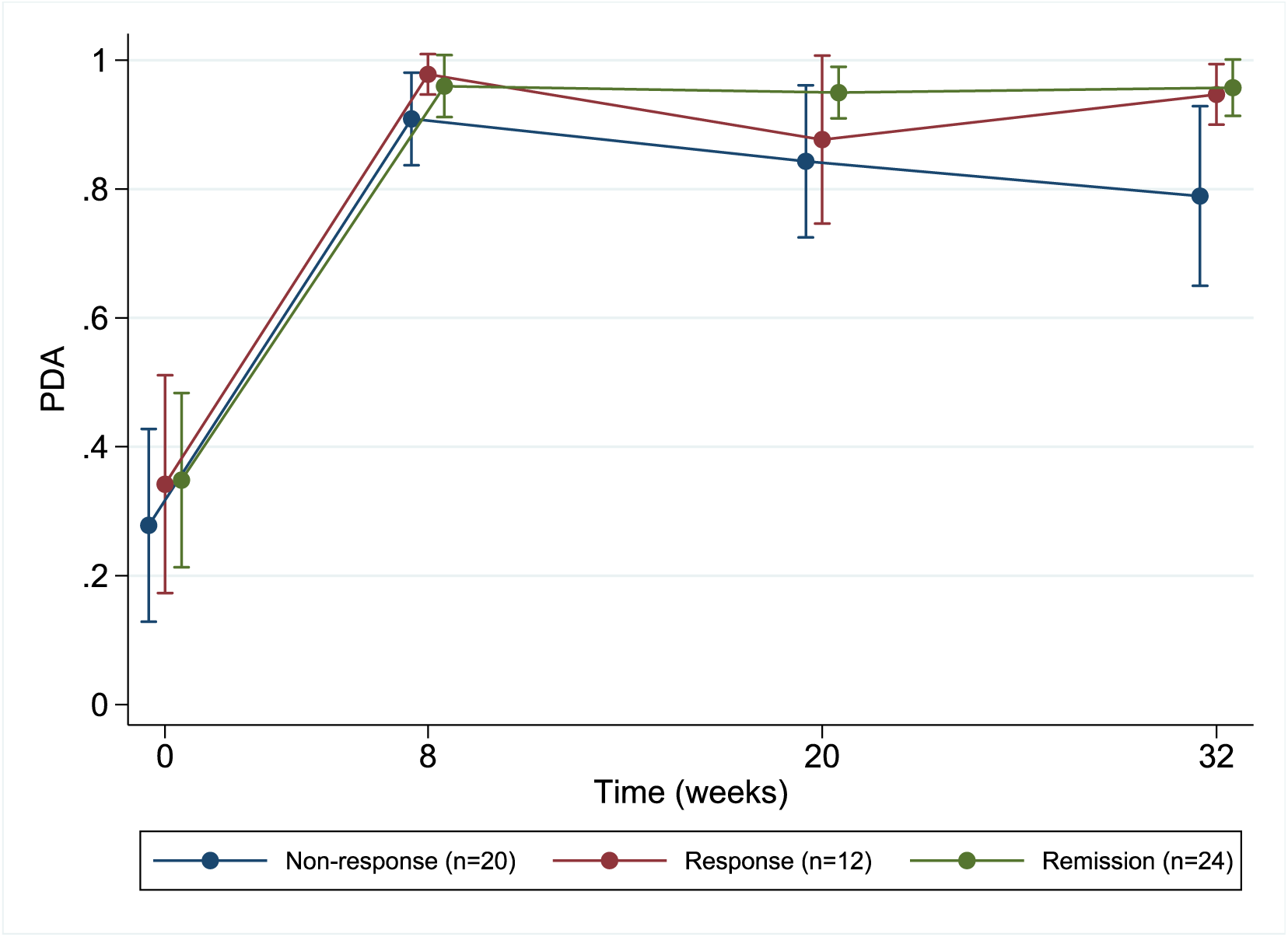
Percent days abstinent (PDA) in the groups based on insomnia treatment outcomes. *Non-response subgroup* (N = 20) included those with neither response nor remission; *Response subgroup* (N = 12) included treatments responders, i.e., those with a per-post change in ISI<7; *remission subgroup* (N = 24) included subjects with an ISI total score <8 at week 8. Subjects with missing data who were excluded (N = 7). The results showed that there was no difference in the PDA between the subgroups over time (p=0.65).

## Notes

### Clinical Trial

NCT01987089

### Author Declarations

IRB of Cpl. Michael J. Crescenz Veterans Affairs Medical Center gave ethical approval for this work

